# SARS-CoV-2 diversity and transmission on a university campus across two academic years during the pandemic

**DOI:** 10.1101/2024.02.29.24303285

**Authors:** AM Casto, MI Paredes, JC Bennett, KG Luiten, J O’Hanlon, PD Han, LS Gamboa, E McDermot, M Truong, GS Gottlieb, Z Acker, CR Wolf, A Magedson, NK Lo, D McDonald, TC Wright, KM McCaffrey, MD Figgins, JA Englund, M Boeckh, CM Lockwood, DA Nickerson, J Shendure, TM Uyeki, LM Starita, T Bedford, HY Chu, AA Weil

## Abstract

Institutions of higher education (IHEs) have been a focus of SARS-CoV-2 transmission studies but there is limited information on how viral diversity and transmission at IHEs changed as the pandemic progressed. Here we analyze 3606 viral genomes from unique COVID-19 episodes collected at a public university in Seattle, Washington (WA) from September 2020 to September 2022. Across the study period, we found evidence of frequent viral transmission among university affiliates with 60% (n=2153) of viral genomes from campus specimens genetically identical to at least one other campus specimen. Moreover, viruses from students were observed in transmission clusters at a higher frequency than in the overall dataset while viruses from symptomatic infections were observed in transmission clusters at a lower frequency. Though only a small percentage of community viruses were identified as possible descendants of viruses isolated in university study specimens, phylodynamic modelling suggested a high rate of transmission events from campus into the local community, particularly during the 2021-2022 academic year. We conclude that viral transmission was common within the university population throughout the study period but that not all university affiliates were equally likely to be involved. In addition, the transmission rate from campus into the surrounding community may have increased during the second year of the study, possibly due to return to in-person instruction.

## Introduction

The Covid-19 pandemic was marked by serial emergence of new SARS-CoV-2 variants which outcompeted and replaced older variants in the general population^1^. Geographical and chronologic variation in circulating variants was revealed through viral genome sequencing surveillance^2^. These efforts were integral for identifying functional differences among variants with regards to transmissibility, symptom profile and severity, and degree of neutralization by antibodies resulting from previous infection or vaccination^3^. Genomic surveillance also helped to predict fluctuations in incidence of COVID-19 and has aided public health authorities in devising mitigation strategies and informing new vaccine formulations and recommendations^4^.

Educational institutions, including institutions of higher education (IHE), have been a major focus of surveillance efforts during the pandemic. These efforts have been aimed at improving understanding of transmission and informing infection control strategies. Though IHEs vary widely in size, demographics, and setting, most IHE populations are predominantly made up of young, healthy adults, that are at relatively low risk of severe disease from SARS-CoV-2^5,6^. Risk may be further mitigated by high rates of vaccine uptake in the setting of vaccine mandates at some IHEs^7^. However, features of IHE environments, such as communal housing and frequent social events, may promote the spread of respiratory viruses, including SARS-CoV-2^8,9^. IHE populations tend to be highly mobile, with students travelling to domestic or international locations during academic breaks, which may contribute to the dispersal of new variants^9^.

SARS-CoV-2 epidemiology and transmission studies on IHE campuses have estimated varying viral transmission rates between IHE populations and their surrounding communities^8,10–15^. Some of this variation is likely due to fixed differences among IHEs, as well as temporal variation in viral transmission dynamics across the pandemic. Most studies of SARS-CoV-2 in IHE populations to date have analyzed data collected during relatively short time frames (such as single academic quarters or semesters), preventing an accurate assessment of how transmission changed over time and how new viral variants and changes in mitigation strategies and human behavior shaped transmission as the pandemic progressed.

Here we examine SARS-CoV-2 genomic sequence data and demographic, epidemiologic, and clinical data collected across from September 2020 to September 2022 as part of a university testing program, the Husky Coronavirus Testing (HCT) study. We used three different means to assess viral diversity and transmission. First, we characterized diversity of viruses seen on campus and compared this to viral diversity within the state. Second, we identified clusters of closely related HCT sequences and assessed the impact of demographic, epidemiologic, and clinical factors of study participants on cluster membership. Third, we examined phylogenetic relationships between university and community viruses.

## Methods and Materials

### Study Overview

Data analyzed in this study were collected as part of the Husky Coronavirus Testing (HCT) study. HCT provided SARS-CoV-2 testing for affiliates (students, faculty, and staff) for the University of Washington (UW) main campus and two satellite campuses from September 2020 to July 2023. Details of specimen and demographic, epidemiologic, and clinical data collection are described in prior studies^16,17^. Briefly, participation was open to all English-speaking university affiliates. Participants completed an electronic questionnaire at enrollment and were sent daily attestation surveys. Throughout the study, participants were invited to test if they reported new symptoms, have a known exposure to a SARS-COV-2 case, or were members of a group experiencing an outbreak. Walk-in testing was also available for any reason. In addition, participants were invited to test from September 2020 to August 2021 following attendance at gatherings with >10 people, from September 2021 to July 2022 following out of state travel, and from August – September 2022 following report of a positive rapid test. The UW IRB approved this study (#00011148). All participants gave informed consent or assent and parent/guardian consent for participants under 18 years of age.

### Swab collection

SARS-CoV-2 testing was performed via (1) participant self-swab collection observed by study personnel at testing sites on campus, (2) unobserved self-swab collection returned to an on-campus drop box, and (3) unobserved self-swab collection picked up by courier. Two swab types were used for observed swab collection at kiosks; US Cotton #3 Steripack Polyester Spun Swabs placed in a 10 mL tube were used at the beginning of the study with transition to RHINOsticTM RH_S000001 Automated Nasal Swabs placed in a MatrixTM 1.0 mL Thermo Fisher 3741 ScrewTop tube in 2021. Swabs returned to drop boxes were RHINOsticTM while those returned via courier were US Cotton #3 swabs.

### Specimen testing for SARS-CoV-2

Specimens collected prior to November 18, 2020 were placed in universal transport media and viral nucleic acid was extracted using two commercial kits as described previously^18^. Specimens collected after November 18, 2020 were stored without preservatives or media. Specimens were prepped for testing using an extraction-free protocol^18^. Aliquots of 5µL were used in four multiplexed RT-qPCR reactions. Two reactions used custom probe sets that targeted Orf1b and two used probes targeted the S-gene. Viral probe sets were multiplexed with a probe set for human RNase P. Specimens were positive for SARS-CoV-2 if viral gene targets and human RNase P were detected in at least three of four reactions.

### Genome Sequencing

Genome sequencing was attempted on all specimens that tested positive for the presence of SARS-CoV-2 with an average cycle threshold of 30 or less. Magna Pure 96 kits (Roche) were used to extract nucleic acids from specimens and sequencing libraries were prepared using COVIDSeq kits (Illumina). Sequencing primers were updated at several points during the study period to account for emergence of new viral variants. Sequencing was performed using NextSeq2000 P200 kits (Illumina). Processing of raw sequence data and generation of consensus genomes was performed using a publicly available bioinformatic pipeline (https://github.com/seattleflu/assembly). All genome sequences used in this study were submitted to Global Initiative on Sharing All Influenza Data (GISAID)^19^.

### Genomic Analyses

SARS-CoV-2 genomes used in analyses that were generated outside of the HCT study were downloaded from GISAID. Both HCT and GISAID genomes were screened for quality using Nextclade CLI^20^. Nextclade was also used to assign sequences to Nextstrain clades and Pango lineages. Sequences given an unfavorable quality rating by Nextclade (based on a sequence’s complement of missing data, mixed sites, private mutations, mutation clusters, frameshifts, and premature stop codons), with a missing collection date, or for which Nextclade was unable to make a clade and/or lineage assignment were excluded from further analysis. Sequence alignment, masking of problematic loci, and phylogenetic tree generation were performed using Nextstrain^21^. Trees were visualized using Auspice. Groups of identical SARS-CoV-2 sequences were identified using a previously described R package (https://github.com/blab/size-genetic-clusters)^22^. Each group of identical sequences were characterized by a set of mutations relative to the reference genome (Wuhan/Hu-1/2019, GenBank Accession MN908947). Any genome that carried the same mutations as a group of identical genomes plus additional mutations relative to the reference was categorized as a descendant of that group. Phylogenetic groups were identified as follows: a phylogenetic tree was constructed for each Nextstrain clade, which included all HCT and GISAID sequences from Washington State (WA) belonging to that clade during the study period. All terminal nodes in these trees were designated as HCT or non-HCT. Augur trait was used to assign HCT versus non-HCT states for all internal nodes and to provide a likelihood of each state assignment^23^. All HCT terminal nodes that descended from the same internal node assigned a state of HCT with a likelihood of 95% or greater were grouped into a single phylogenetic cluster.

### Transmission Modeling

Our transmission modeling analysis included SARS-CoV-2 sequences divided into 3 regions of origin: HCT, consisting of HCT sequences; KC, consisting of sequences from King County (KC), WA; and other, which consisted of contextual sequences from around the world to account for outside viral introductions. We employed an equal temporal subsampling scheme to enrich for under sampled time periods by randomly choosing a maximum of 400 total sequences per region (HCT, KC, and other) sampled equally per each calendar month via Augur filter^23^, resulting in a set of 1137 total sequences, which were input into the model. Given the differential number of specimens in each region-year-month combination, not all demes included a total number of 400 sequences. We chose an equal temporal subsampling scheme based on recent work showing that maximizing spatiotemporal diversity reduces bias in MASCOT^24^.

Using the compiled input sequence set, we employed a MASCOT-Skyline approach, which approximates the structured coalescent, to predict when the most recent common ancestor for each sequence pair in our input set existed and which of the three regions this ancestor would have existed in. To generate these predictions, we made assumptions about effective population sizes of the three regions and migration among the regions. To allow for population sizes to change over time, we modeled effective population sizes similar to the Skygrid approach for unstructured populations^25^. We estimated the effective population size for each location between time *t*=0×tree height, …, *t*=1×tree height. Between each time point where we estimated *Ne*, we assumed exponential growth. *A priori*, we assumed that the effective population size at time *t*+1 is normally distributed with mean 0 and standard deviation α, with α being estimated. We assumed the migration rate to be constant forward-in-time, 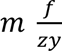 , between states *y* and *z*. As the structured coalescent assumes backwards-in-time migration rates, we assumed that backwards-in-time rate of migration between state *y* and z, 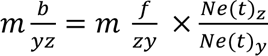. To infer effective population sizes and migration rates over time, we employed an adaptable multivariate gaussian operator^26^.

Parameter traces were visually evaluated for convergence using Tracer (v1.7.1)^27^ and 30% burn-in was applied for all phylodynamic analyses. Output from our modeling analysis was a phylogenetic tree with internal nodes representing common ancestors of input sequence pairs. Tree plotting was performed with baltic (https://github.com/evogytis/baltic) and data visualizations were done using Altair^28^. We summarized trees as maximum clade credibility trees using TreeAnnotator and visually inspected posterior tree distributions using IcyTree^29^. Transmission between regions was calculated by measuring the number of migration jumps from HCT to KC and vice versa walking from tips to root in the posterior set of trees. Persistence time was measured by calculating the average number of days for a tip to leave its sampled location (HCT, KC, other), walking backwards up the phylogeny from tip up until node location was different from tip location^30^.

## Results

### Viral lineages and clades common in Washington State were observed among HCT specimens

We sequenced 3,855 of 6,485 SARS-CoV-2 positive specimens collected by HCT from September 2020 to September 2022. These sequences represent 3% of all SARS-CoV-2 genomes generated from specimens collected in Washington State (WA) during this time. From this raw sequence set, we retained only one sequence per person per infection and filtered out poor quality sequences, resulting in 3,606 sequences (Figure 1) in the final dataset; 3195 of these sequences were collected during academic year 2 (September 1, 2021 – September 30, 2022; hereafter referred to as year 2) with 1813 collected between December 1, 2021 and February 28, 2022 (Supplementary Figure S1). The final sequence set contained sequences from 19 different Nextstrain clades and 115 Pango lineages (Supplementary Note S1; Supplementary Tables S1, S2).

**Figure 1:**
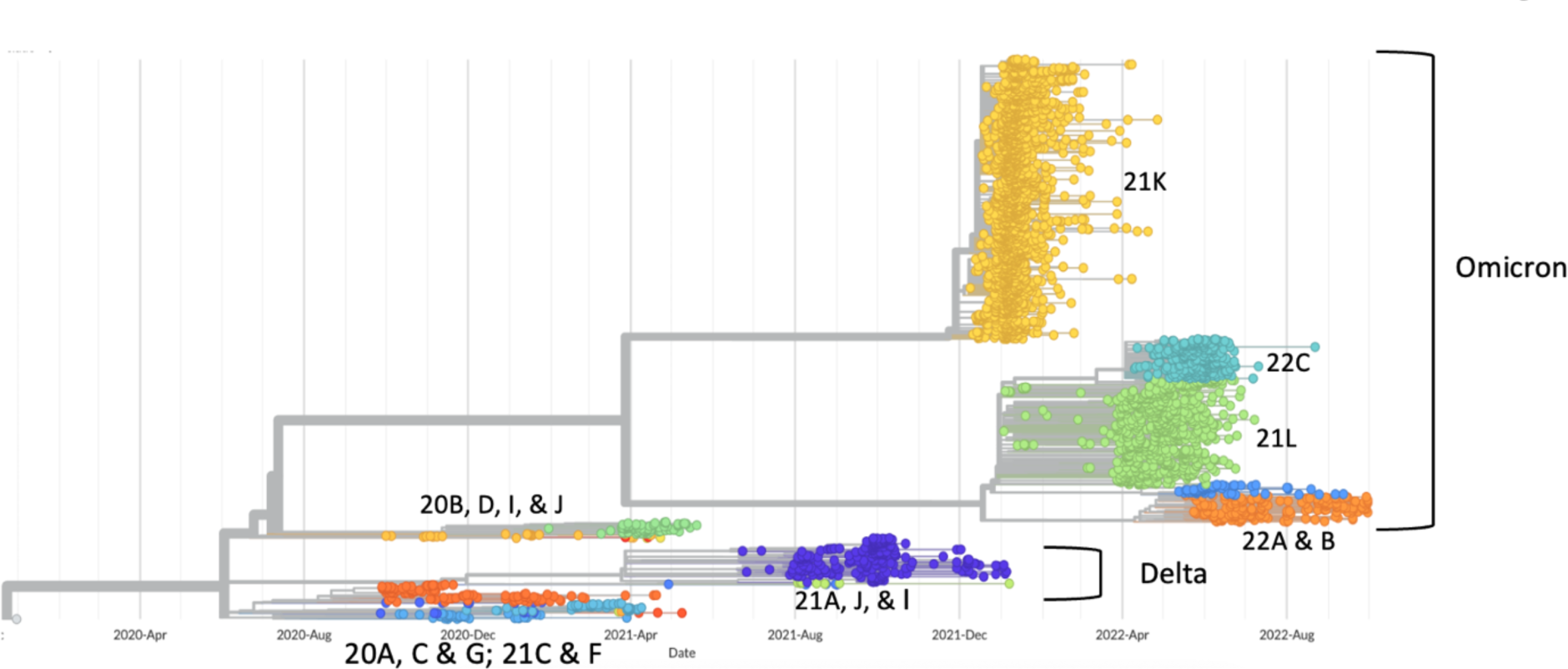
Phylogenetic tree of all 3606 HCT sequences. Tips are colored by Nextstrain clade. Date of specimen collection is on the x-axis.

To provide context for diversity seen among SARS-CoV-2 genomes from HCT specimens, we downloaded all SARS-CoV-2 genomes from specimens collected in WA outside of the HCT study from September 2020 to September 2022 from GISAID EpiCoV database^19^. After filtering out poor quality and duplicate sequences, a total of 119,215 WA genomes remained, representing 27 different clades and 333 lineages. All clades with a frequency of >0.2% and all lineages with a frequency of >0.4% among WA genomes were represented by at least one HCT genome. Most lineages in WA were rare (<0.4% of all WA genomes) and so more than half (n=224, 67.3%) of all WA lineages were not observed among HCT genomes. There were 6 lineages that were represented in HCT but not the WA sequence set. These were all from samples collected in early January or late March 2022 (Supplementary Table S3). The percent of WA clades and lineages observed in HCT fluctuated over time; in year 2, these percentages appeared to spike at the beginning of academic quarters (Supplementary Figure S2; Supplementary Table S4).

### Average delay of one month between variant observation in WA and in HCT

The prevalence of clades among HCT and WA sequences over time is shown in Figure 2. For clades and lineages seen among both HCT and WA genomes, we determined the date of first observation of a lineage and clade in each group. The average number of days from observation in WA to observation among HCT specimens was 35.1 days (median 24, range 0 – 116) for clades and 35.5 days (median 28, range −56 to 170) for lineages (Supplementary Figure S3). Ten lineages were observed among HCT specimens before WA specimens. Notably, the BA.2 lineage, which was represented by 428 (11.9%) HCT sequences and 6,704 (5.6%) WA sequences and from which all currently circulating SARS-CoV-2 are descended, was among these and was first observed on campus on January 3, 2022. Three of the other lineages first observed in HCT were also collected in January 2022.

**Figure 2:**
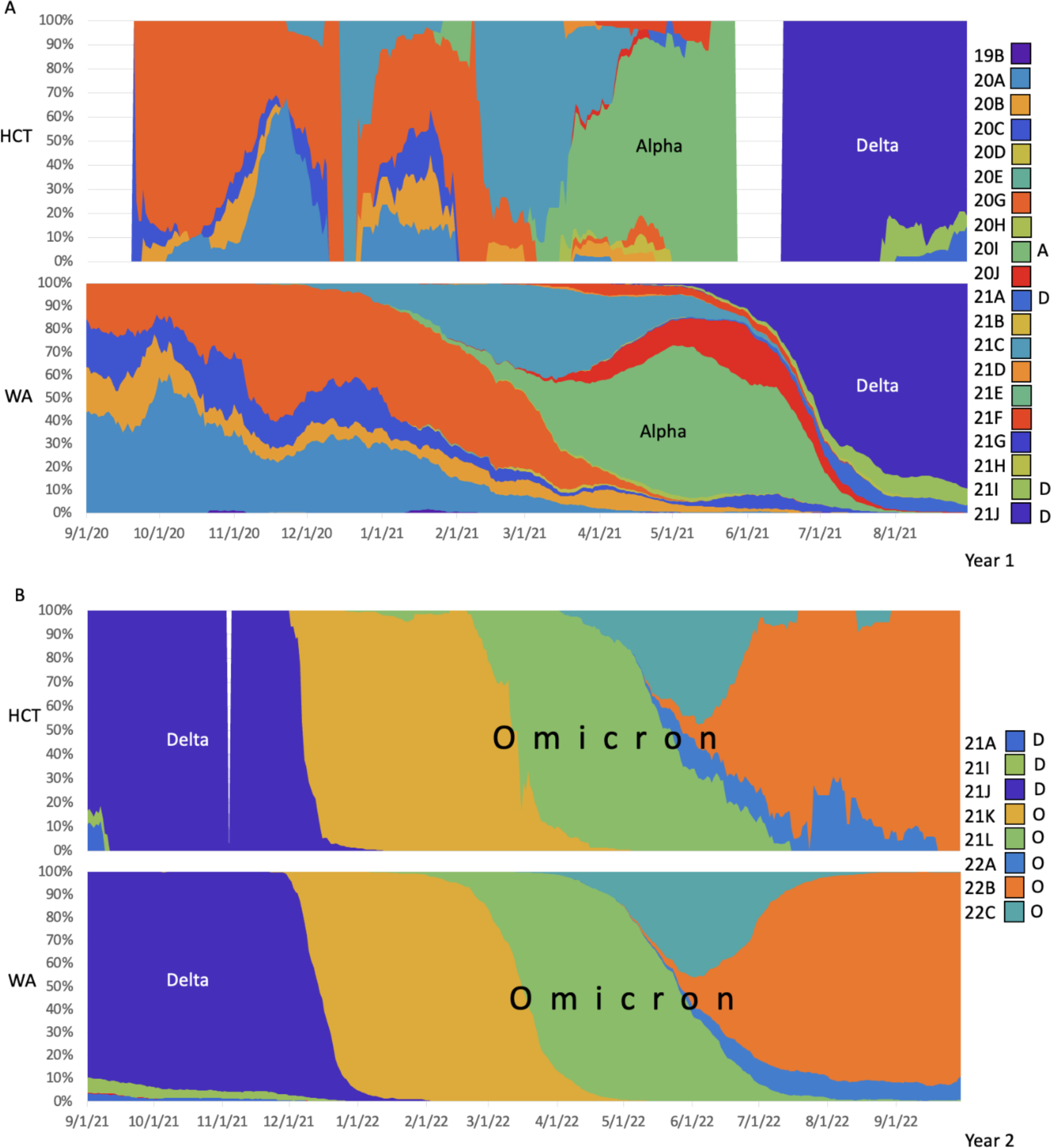
Frequency of Nextstrain clades among HCT and WA SARS-CoV-2 genomes over time. Each time point on the x-axis represents a two-week sliding window centered on that date. Y-axis shows the distribution of genomes collected in that window among different clades. Blank sections for HCT represent two-week windows during which no sequenced specimens were collected. Chart colors correspond to Nextstrain clades as shown in the legend. Alpha variant clades are labeled “A”, Delta variant clades are labeled “D”, and Omicron variant clades are labeled “O” in the legend. A) Specimens collected during year 1 (September 1, 2020 to August 31, 2021). B) Specimens collected during year 2 (September 1, 2021 to September 30, 2022).

### Most HCT SARS-CoV-2 specimens were closely related to at least one other HCT specimen

We used two different approaches to identify groups of closely related HCT SARS-CoV-2 genomes. First, we identified groups of identical genomes (which we refer to as “zero distance clusters”). There were 1730 unique haplotypes among HCT sequences, including 2153 sequences that were identical to at least one other HCT sequence and 277 haplotypes represented by more than one HCT sequence (Figure 3A, Supplementary Table S5, Supplementary Figure S4). A single Omicron haplotype (clade 21K, lineage BA.1.1) was observed for 655 different sequenced specimens collected from December 17, 2021 until March 8, 2022 (18.2% of all HCT genomes). Of the 277 zero distance clusters, 26 included 10 or more sequences. For each clade, the average size of zero distance clusters decreased with time since clade introduction (Supplementary Figure S5)^22^ consistent with declining transmission rate over time following variant introduction. The longest period over which a single haplotype was observed was 153 days (clade 21L, lineage BA.2).

**Figure 3:**
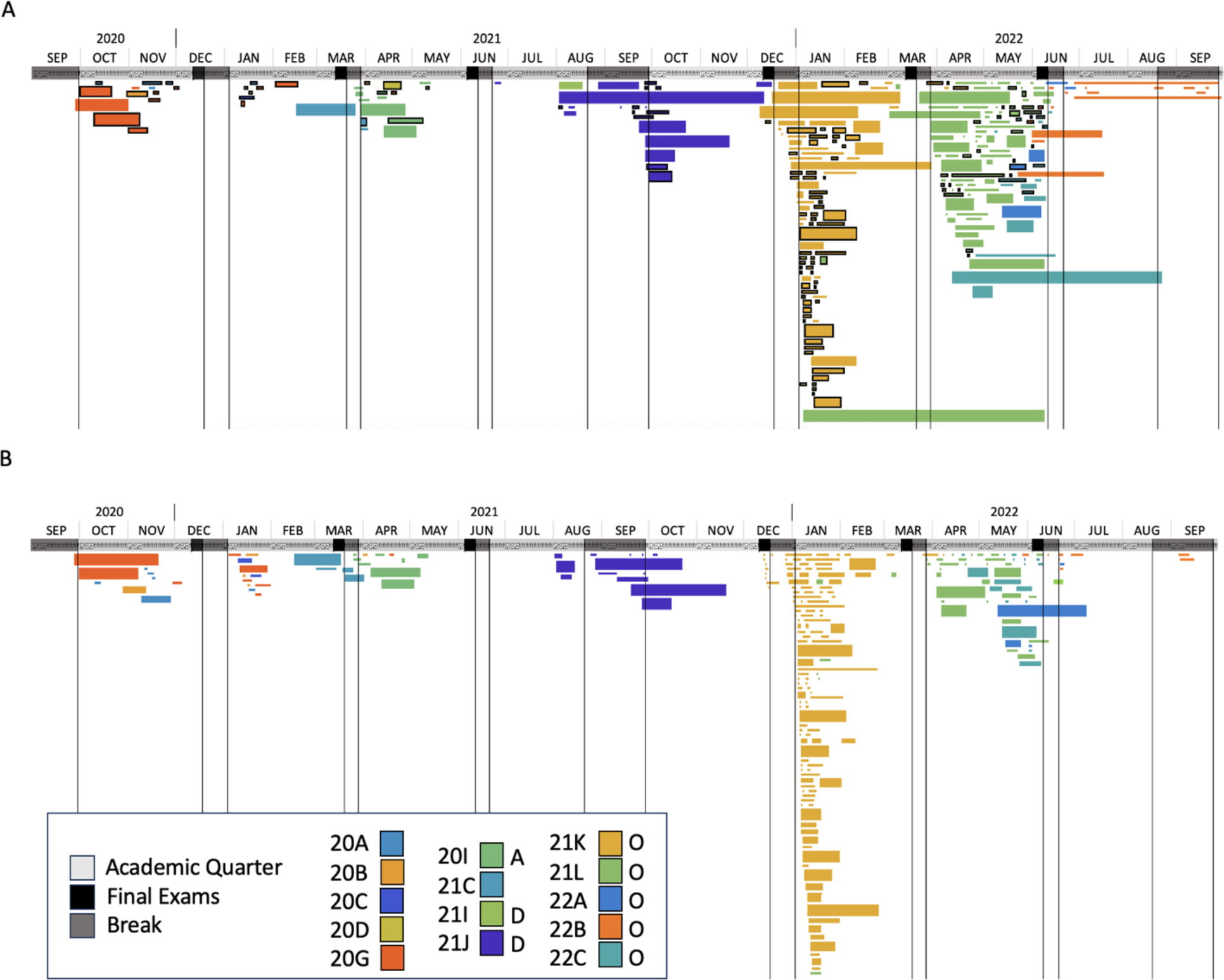
Chronology of clusters of HCT sequences. Each bar represents a sequence cluster which extends from the collection date of the first specimen in the cluster to the collection date of the last specimen. Month and year of specimen collection is denoted at the top with dates of academic quarters in light gray, final exams in black, and university breaks in dark gray. Color of cluster bars corresponds to viral clade as indicated in the legend. Alpha variant clades are labeled “A”, Delta variant clades are labeled “D”, and Omicron variant clades are labeled “O” in the legend. Height of cluster bar is proportional to the number of sequences in the cluster. A) Zero distance clusters (groups of identical sequences). Bars with black outlines represent HCT-only zero distance clusters (groups of identical sequences with haplotype unique to HCT). B) Phylogenetic clusters (groups of sequences that cluster phylogenetically).

We also identified groups of identical sequences among a combined HCT and WA dataset. Of 277 HCT zero distance clusters, 133 (48%) represented a haplotype not observed among WA genomes (we refer to these as “HCT-only zero distance clusters”). The largest HCT-only zero distance cluster (clade 21K, BA.1.20) included 12 sequences and the most persistent cluster (longest period from collection of first to last specimen) was observed over a period of 35 days (clade 21K, BA.1.1). To assess for possible “spill-over” of virus from university affiliates into other populations, we looked for WA viruses that appeared to be descendants of one of 133 HCT-only zero distance clusters (see Methods). We found a total of 81 such non-HCT viruses, associated with 19 clusters (Supplementary Tables S6, S7). Over half (n=42, 51.9%) of these 81 viruses were of the BA.2 lineage (clade 21L). The largest number of non-HCT descendants of a single cluster was 37 (clade 21L, BA.2, Supplementary Figure S6).

We created a phylogenetic tree for each clade that included all HCT and WA genomes. We used these trees to identify clusters (which we refer to as “phylogenetic clusters”) of HCT genomes that descend from a single introduction event (see Methods). These clusters ranged in size from 2 to 70 sequences with 19 clusters including more than 10 sequenced specimens (Figure 3, Supplementary Table S8). Most (n=198, 84.6%) of the 234 HCT phylogenetic clusters included only HCT sequences. However, a total of 218 WA sequences were part of an HCT sequence cluster. The largest number of non-HCT sequences in a single HCT phylogenetic cluster was 37 (clade 20I, lineage B.1.1.7; Supplementary Tables S9, S10).

### Model suggests high transmission rate from the university into the surrounding community

To further explore the relationship between SARS-CoV-2 in university affiliates and the surrounding community, we modeled transmission dynamics to and from the HCT population. We limited the WA sequences in this analysis to those from King County (KC) to more accurately reflect the community immediately surrounding the university. In addition to including an HCT and KC region in the model, we also included an “other” region representing sequences from outside KC in WA and the rest of the world. After subsampling (see methods), a total of 1137 genomes were used as input for the model. Results suggested a higher forward migration rate from HCT into KC than vice versa (Figure 4; 10.8 migration events/lineage/year [95% highest posterior density (HPD) 4.3-19.9] vs 0.13 migration events/lineage/year [95% HPD 0.068-0.179]). We estimated that KC had at least 433 (IQR: 415-444) viral introduction events during the study period with at least 130 events (IQR: 126 – 137) coming from the HCT population. These numbers represent the lower bound of the number of introductions as the absolute number is constrained by the total number of sequences in our specimen set. Our model indicated that viral lineages are more likely to circulate longer in the larger KC region (92.6 days, IQR: 86.4-101.1 days) than in the HCT population (77.2 days, IQR: 71.5 – 82.6 days). When analyzing transmission patterns across time, we find that viral flow between HCT and KC was dominated by spread from KC to HCT during academic year 1 (September 1, 2020 – August 31, 2021, hereafter referred to as year 1), and from HCT to KC during year 2 (Figure 5).

**Figure 4:**
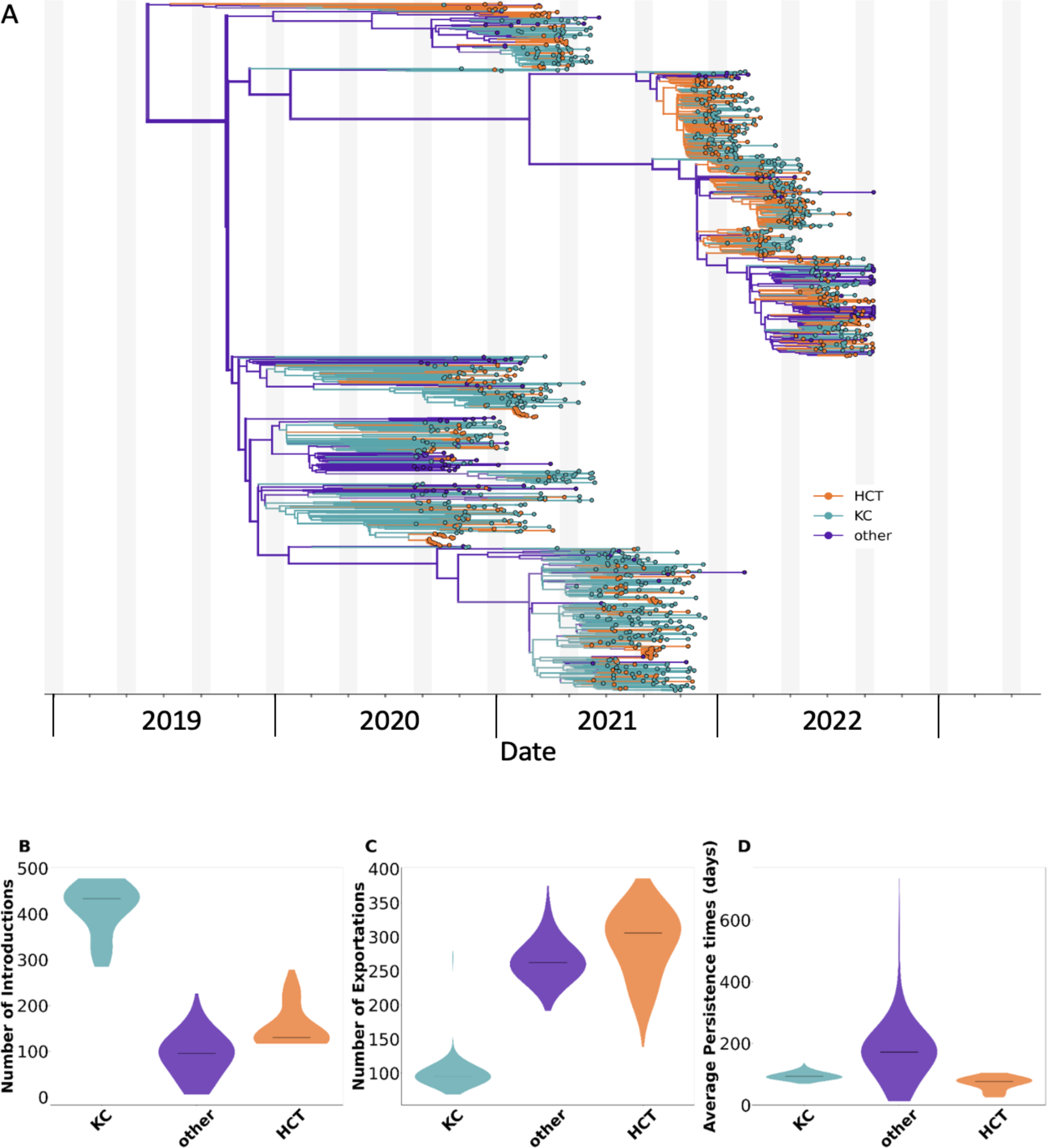
Phylodynamic analysis of SARS-CoV-2 transmission between KC and HCT populations. A) Maximum clade credibility tree summary of the Bayesian inference conducted using MASCOT-Skyline on 1137 sequences. Colors correspond to the locations in the legend. KC = Sequences from King County (excluding those from HCT), HCT = Sequences from specimens collected by HCT, other = global contextual sequences from outside of King County sampled to increase spatiotemporal diversity. Estimated number of introductions B), exports C), and average time of local persistence in days D) for each region. Horizontal black line denotes median estimates.

**Figure 5:**
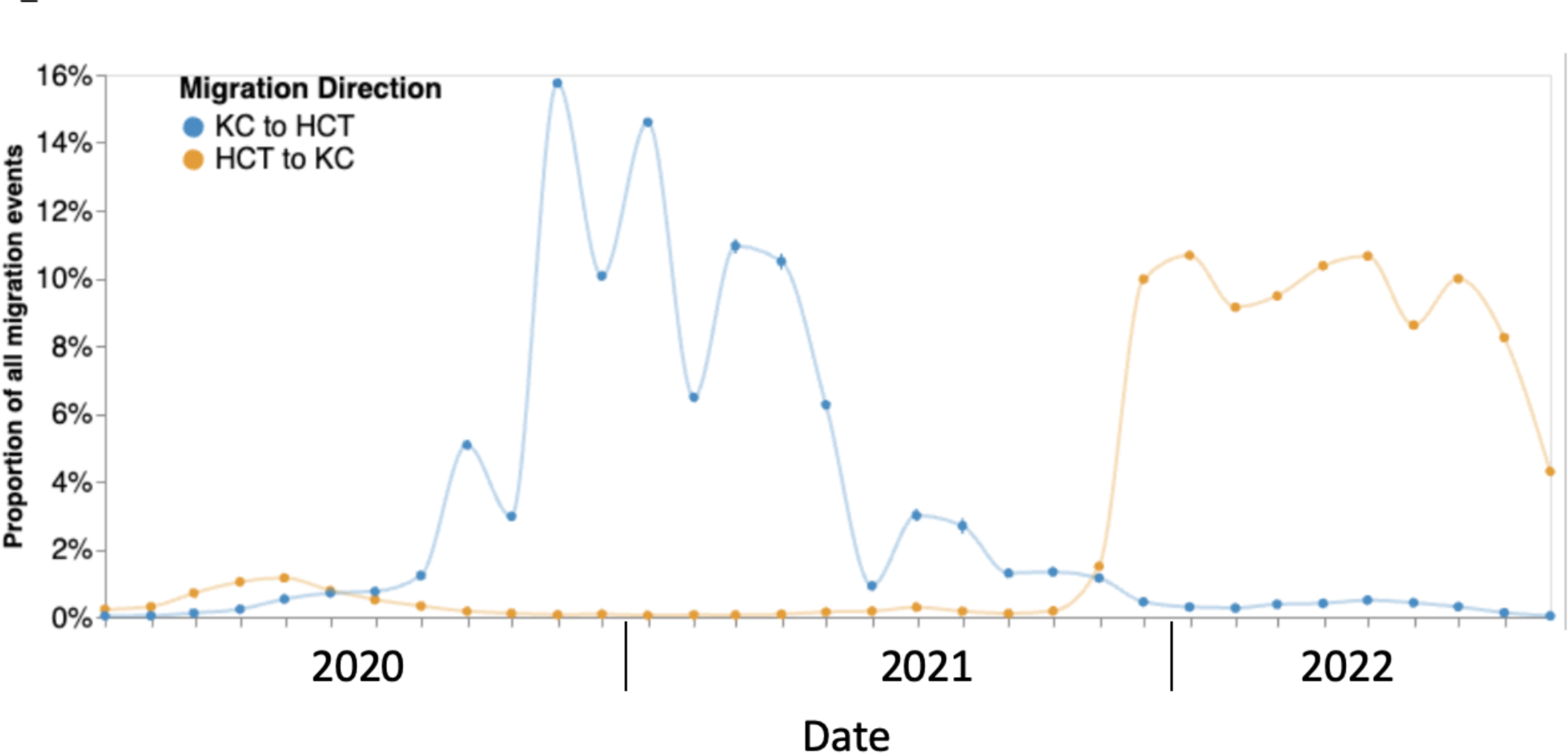
Proportion of all migration events between HCT and KC. KC to HCT migration is in blue and HCT to KC migration is in orange. Proportions do not add up to 100% as migration events including the “other” region were excluded.

### Participants with a sequenced viral genome were representative of those testing positive for SARS-CoV-2

The 3,606 HCT genomes were from 3,560 unique individuals (Supplementary Table S11). Most (85.4%) were students, 57.5% identified as female, 8.6% were Latinx, and the majority were White (50.4%) or Asian (32.0%). Average age at the time of infection was 25.1 years (median 21.3 years, range 17.4 – 78.7). HCT participants with a sequenced specimen were overall demographically representative of all HCT participants with a positive test (Supplementary Table S12). 3,514 individuals had only one sequence in the dataset while 46 individuals, who experienced infection with more than one clade/lineage of SARS-CoV-2 during the study period, had two sequences included in the dataset (Supplementary Table S13, Supplementary Note S2). Of these, 36 were first infected by a non-Omicron SARS-CoV-2 variant followed by an Omicron variant, 9 were infected by two different Omicron variants, and one was infected with two different non-Omicron variants (Supplementary Note S3).

### Sequences from students, younger persons more likely to cluster with other HCT sequences

We examined the impact of epidemiologic, demographic, and clinical factors of infected persons on the relationship among viruses in the HCT population. Students were overrepresented among re-infected individuals relative to their frequency in the complete dataset (one proportion z-test, 95.7% versus 85.4%, p = 0.048), while symptomatic infections were underrepresented (47.8% versus 75.5%, p < 0.0001, Figure 6, Supplementary Table S14). Additionally, average age at the time of infection was lower for those who experienced re-infection than for all participants with a sequenced virus (21.1 versus 25.1, p < 0.0001). This was also observed when sequences from students and those from faculty/staff were considered separately (20.5 versus 21.7, p < 0.0001 and 33.0 versus 44.8, p = 0.0096).

**Figure 6:**
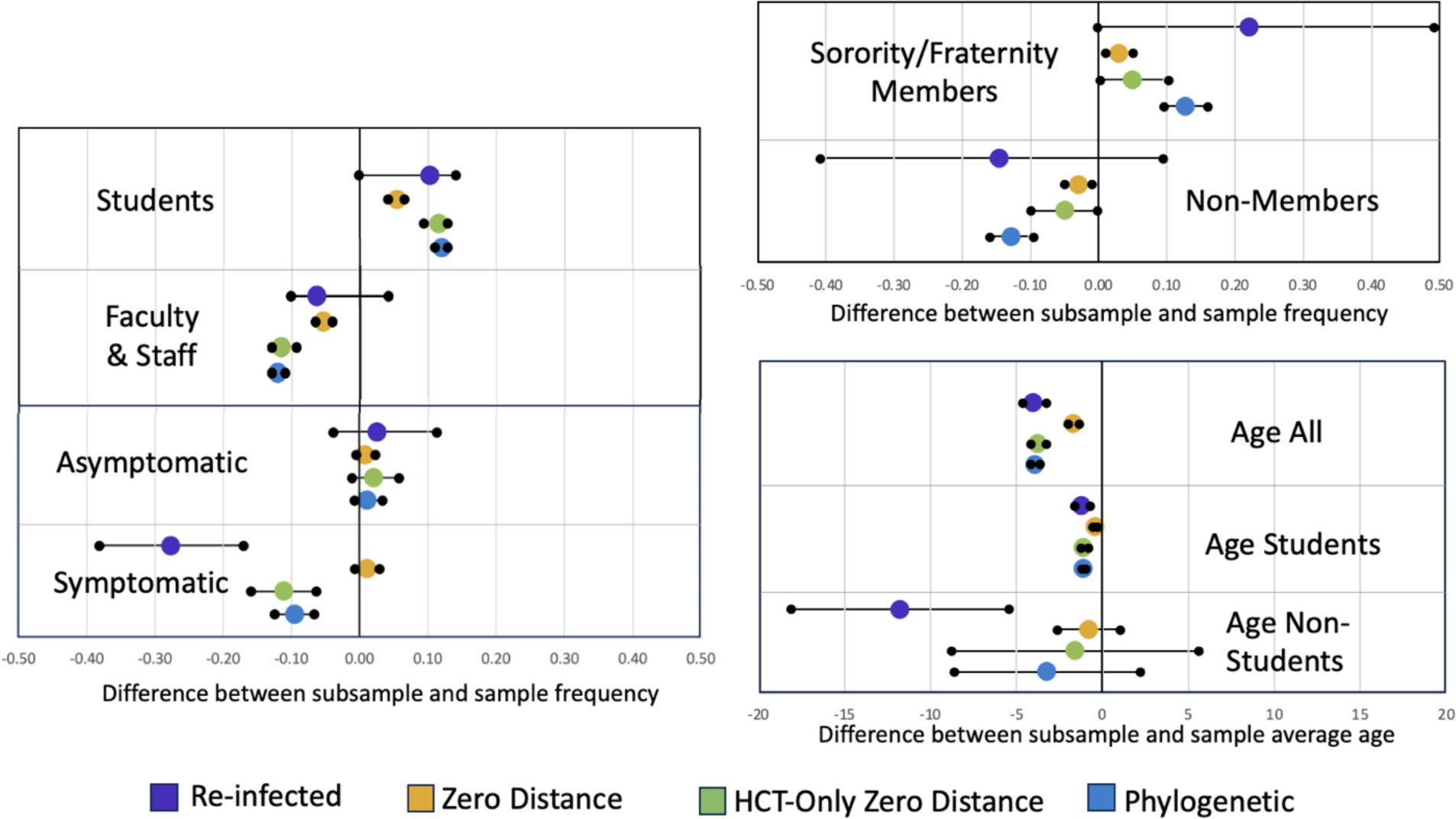
Demographic and epidemiologic characteristics of participants who experienced re-infection and of those with sequenced specimens in clusters. For student, faculty & staff, asymptomatic, symptomatic, sorority/fraternity members, and non-members categories, the mid-line (x = 0) represents the percent representation of that category in the whole dataset. The purple, yellow, green, and blue dots give the percent difference between the frequency of that category among those who experienced re-infection and those with sequences in zero distance (groups of identical sequences), HCT-only zero distance (groups of identical sequences with haplotype unique to HCT), and phylogenetic clusters (groups of sequences that cluster phylogenetically), respectively, and its frequency in the whole dataset. Intervals marked by black dots connected by a bar indicate the 95% confidence interval. For age all, age students, and age non-students categories, the mid-line (x = 0) represents the average age at time of infection for all sequences, sequences from students, and sequences from non-students, respectively. The purple, yellow, green, and blue dots give the difference between the above averages and the average age of those who experienced re-infection and for sequences in zero distance, HCT-only zero distance, and phylogenetic clusters, respectively. Intervals marked by black dots connected by a bar indicate the 95% confidence interval.

Sequences from students were overrepresented among those in zero distance, HCT-only zero distance, and phylogenetic clusters (90.9%, 97.0%, 97.6% versus 85.5%, p < 0.0001 for all) while sequences from non-students (faculty/staff/other) were underrepresented in all three cluster types (9.1%, 3.0%, 2.4% versus 14.5%, p < 0.0001 for all). Sorority/fraternity members were also overrepresented in all three cluster types (21.5%, 23.5%, 31.3% versus 18.6%, p = 0.00022, 0.0276, <0.0001), while sequences from symptomatic infections were underrepresented among those in HCT-only zero distance and phylogenetic clusters (64.4%, 65.9% versus 75.5%, p < 0.0001 for both). Finally, average age at the time of infection was lower for sequences in all three cluster types compared to average age for all sequences (23.4, 21.3, 21.1 versus 25.1, p < 0.0001 for all). This difference was also observed when sequences from students were considered separately (21.3, 20.6, 20.6 versus 21.7, p < 0.0001 for all). Average ages at infection for sequences from non-students in all 3 cluster types did not differ from the overall average age for non-students (44.0, 39.2, 43.2 versus 44.8, p = 0.3922, 0.6292, and 0.2305). Results of similar analyses for other demographic and epidemiologic variables are shown in Supplementary Figure S8.

## Discussion

We studied SARS-CoV-2 cases with associated viral genomic data in a large, public university population over the first two academic years of the pandemic with a focus on characterizing viral diversity and transmission dynamics. To our knowledge, this represents the largest survey to date of IHE SARS-CoV-2 cases with viral sequence data and one of the few based on data collected for more than one year. Our results provide an in-depth analysis of SARS-CoV-2 in an IHE population during many changes in viral epidemiology, transmission mitigation strategies, and human behavior. We found that some measures of viral diversity and transmission dynamics differed between year 1, during which online-only instruction occurred, and year 2, during which classes were conducted mostly in-person, or appeared to be impacted by the academic calendar; other measures were stable throughout the study period. We also observed that not all university affiliates were equally likely to be involved in campus-related viral transmission. Students and sorority/fraternity members were overrepresented in transmission clusters while faculty/staff and those with symptomatic infections were under-represented relative to their representation in the full dataset. In addition, the average age at the time of infection of those in transmission clusters was lower than the overall average age and the average age of students in clusters was lower than the average age for all students. These findings provide context for and aid in interpretation of other studies of SARS-CoV-2 at IHEs, particularly those with shorter study periods. They also can help administrators of IHEs target mitigation strategies toward affiliates at highest risk of involvement in campus-related SARS-CoV-2 transmission.

Our study has several unique features, including evaluation of viral diversity on campus compared to the state over time. We found that clades/lineages common in the state were reliably observed within HCT even during year 1, when the number of samples collected was relatively small. There was a notable difference between year 1 and year 2 in the similarity between clade frequency on campus and in the state, with HCT clade frequencies observed during study year two almost identical to statewide frequencies. We also noted that the average delay between first observation of a clade or lineage on campus relative to first observation in the state was shorter during year 2 relative to year 1. These differences may be explained by differences in sample size, though we hypothesize that changes in infection control measures and in the virus, such as a return to in-person instruction, the introduction of more transmissible variants, and overall higher infection rates in year 2, also played a role. We observed evidence that the academic calendar influenced the relationship between viral diversity on campus and in the state. We saw spikes in the percent of WA clades and/or lineages represented on campus at the beginning of academic quarters in year 2. Lineages unique to HCT or seen first in HCT were also mostly collected at the beginning of academic quarters in year 2. These observations are consistent with increased importation of viral diversity into the HCT population when students, 10% of whom are international and 15% of whom are out-of-state residents, were returning to campus for the start of in-person instruction from diverse geographic locations. These patterns were not observed in year 1 of the study when classes were held exclusively online.

We used two different methodologies to define putative campus transmission clusters, one based on genetic distance and one based on phylogenic relationships. This was done to mitigate disadvantages of each method and assess robustness of results. When comparing zero distance and phylogenetic clusters, we noted similarities that are likely to be reflective of campus transmission dynamics. Both methodologies suggested that campus related transmission was common throughout the study period as most specimens were closely related to at least one other HCT specimen. Most clusters were confined to a single academic quarter as expected given drop off in campus population during breaks, though interestingly, there were some exceptions to this. Two phylogenetic clusters persisted through spring break 2021; one large Delta phylogenetic cluster started in early September 2021 and persisted into fall quarter 2021, and several Omicron clusters started in late December 2021 and persisted into winter quarter 2022, suggesting some on-going transmission among university affiliates even during academic breaks. Additionally, both cluster types indicated that campus transmission chains could persist for weeks to months despite frequent lineage replacement within the SARS-CoV-2 population. The number of clusters per academic quarter was stable during year 1 and increased during year 2. This seems to be due to the increase in the number of specimens collected in year 2 relative to year 1, rather than a change in transmission dynamics, as the percent of specimens falling into zero distance, HCT-only zero distance, and phylogenetic clusters remained stable across the study period. Finally, we observed that average cluster size (for both zero distance and phylogenetic clusters) for a particular variant decreased with increasing time since emergence of that variant. This is consistent with prior observations in WA^22^ and is thought to be indicative of a decrease in the effective reproduction number for viral variants the longer they have been circulating in a population.

We also conducted a modelling analysis to assess concordance with our other results. The average number of days that viral lineages circulated on campus was estimated to be 77.2. This was surprisingly high given that longest persistence times of the three cluster types were 153 days (zero distance), 35 days (HCT-only zero distance), and 60 days (phylogenetic), but does provide further support for the hypothesis that viral transmission among university affiliates was common during the study period. Interestingly, model results also suggested that viral flow between KC and HCT was mostly into the campus population during year 1 and then from the campus population during year 2. This could be the result of spread of new viral variants (Delta, Omicron), the return to in-person instruction in year 2 with an increase in campus population relative to year 1, or some combination of these. Given that we found few WA sequences that appeared to descend from HCT clusters, we were surprised that the model estimated a substantial number of transmission events from HCT to KC (estimated minimum of 130) and a significantly higher forward migration rate than from KC to HCT. Most of this difference is likely attributable to the vastly different population sizes of the two regions; KC has population of 2.252 million while HCT enrolled 37,360 participants. If we imagine a SARS-CoV-2 transmission chain starting in KC, we expect a 1.7% chance of that transmission chain jumping into HCT due to differences in population size alone. Conversely, if the transmission chain started in HCT, we estimate a 98.3% chain of this chain infecting an individual in KC. This asymmetry corresponds to a 59.2 fold larger viral migration rate from HCT to KC than vice versa. The fact that this magnitude increase is similar but still less than the 85.7 fold difference in estimated forward migration rates suggests that the difference in migration rates can largely be explained by the difference in population sizes, further augmented by the presence of population structure. In summary, the results of the modeling analysis do not suggest that SARS-CoV-2 cases in the HCT population had a disproportionate impact on KC, but also that nearly all HCT transmission chains resulted in an infection in the KC population.

We used clinical, epidemiologic, and demographic data for HCT participants to assess the relationship of these factors to viral clustering. While findings that viruses from students were disproportionately represented in clusters and that average age of those with viruses in clusters was younger than the population average are not surprising, these results provide evidence to support the direction of limited infection control resources at IHEs to those most likely to be involved in transmission chains. Studies with more detailed information about participant housing, activities, behaviors, and vaccination status could help to further delineate drivers of this association between student status, age and, cluster membership to more strategically target infection control resources. Due to outbreaks associated with sorority/fraternity membership at the study university in 2020^16,31^, the HCT study did collect data on participant involvement with these social groups and our results indicated that sorority/fraternity members were disproportionally represented in transmission clusters. Data on membership in other social groups, such as sport teams or clubs, was not collected by HCT and we are unable to comment on the impact of participation in these activities on involvement in campus-related viral transmission. We note, though, that unlike most social groups, sorority and fraternity members frequently live together in communal housing, which could be a major driver of their risk of involvement in viral transmission chains. Finally, differences in clustering observed for symptomatic versus asymptomatic cases was an unanticipated result. One possible explanation is differences in behavior of the two groups, such as increased social distancing and isolation by symptomatic individuals.

Limitations of our study included incomplete case identification and sampling on the university campus and in WA state during the study period. Additionally, sequence data could not be generated for all HCT cases, particularly those involving specimens with low amounts of viral RNA (high cycle thresholds). University affiliates could also test outside of HCT and sequenced specimens from affiliates collected outside HCT were classified as non-HCT WA sequences. This reflects the broader challenge of the lack of associated demographic and clinical data for most SARS-CoV-2 genomes in GISAID. This limits our understanding of relationships between viral transmission and factors such as gender, age, race/ethnicity, symptoms, and place of residence below the level of state. In addition, changing availability of genomic surveillance data over time and unequal sampling across WA and the world impacted the probability that a case was represented by a sequence in our dataset. We attempted to mitigate this potential bias in our modeling analysis by using spatiotemporal subsampling, which has been shown to improve inferential power of similar models^24^. However, conclusions of our modeling analysis, and all similar modeling analyses, are limited by the fact that results are based on assumptions about population sizes and migration rates, which may be inaccurate, and on the input sequence set, which represents a small fraction of all SARS-CoV-2 cases occurring in the three regions during the study period.

Populations of IHEs have been and will continue to be a focus of SARS-CoV-2 research out of concern that these populations are prone to frequent transmission, which may have significant impacts on IHEs and surrounding communities. Varying results have made it challenging to derive generalizable lessons from studies conducted in IHEs during the last several years. Here we have characterized viral diversity and transmission at a single IHE over two years to gain an understanding of how viral diversity and transmission dynamics at a single institution can vary over time and to aid in the synthesis of the data and results from previous studies into a cohesive knowledge base. Such knowledge is vital to future optimization of interventions to limit spread of SARS-CoV-2 and other respiratory viruses in IHE populations.

## Data Availability

All SARS-CoV-2 genomes used in this study have been deposited to GISAID (https://gisaid.org/).
Additional data and software files are available on github (https://github.com/amcasto/huskytesting_SARSCoV2genomics_First2Years).

https://github.com/amcasto/huskytesting_SARSCoV2genomics_First2Years

## Acknowledgements

We would first like to thank all the study participants and the HCT study team. We would also like to thank the University of Washington, including UW Environment Health and Safety team (Katia Harb, Sheryl Schwartz, Natalie Thiel, Kim Baker, and Julie Skene) and the UW Covid Incident Command team (Margaret Shepherd, Josh Gana, Pamela Schreiber, and Jack Martin). Finally, we would like to thank all contributors of data to GISAID. We have included a GISAID acknowledgements table in the Supplementary Material (Supplementary Table S15).

## Funding

This work was supported by a Howard Hughes Medical Institute Covid Supplement Award to TB and by the United States Senate and House of Representatives, Bill 748, Coronavirus Aid, Relief, and Economic Security Act. MIP is an ARCS Foundation scholar. TB is a Howard Hughes Medical Institute Investigator.

## Data Availability Statement

All SARS-CoV-2 genomes used in this study have been deposited to GISAID (https://gisaid.org/). Additional data and software files are available on github (https://github.com/amcasto/huskytesting_SARSCoV2genomics_First2Years).

## Authorship

Conceptualization: AMC, MIP, TB, HYC, AAW; Methodology: AMC, MIP, TB; Software: AMC, MIP, KGL, TB; Validation: AMC, MIP; Formal Analysis: AMC, MIP; Investigation: JCB, KGL, JO, PDH, LSG, EM, MT, ZA, CRW, AM, NKL, DM, TCW, KMM; Resources: GSG, JAE, MB, CML, DAN, JS, LMS, TB, HYC, AAW; Data Curation: JCB, KGL, JO, PDH, LSG, EM, MT, ZA, CRW, AM, NKL, DM, TCW, KMM; Writing – Original Draft Preparation: AMC, MIP, TB, HYC, AAW; Writing – Review & Editing: All Authors; Visualization: AMC, MIP; Supervision: GSG, JAE, MB, CML, DAN, JS, TMU, LMS, TB, HYC, AAW; Project Administration: JCB, GSG, ZA, CRW, NKL, JAE, MB, CML, DAN, JS, LMS, TB, HYC, AAW; Funding Acquisition: JAE, MB, CML, DAN, JS, LMS, TB, HYC, AAW

## Competing Interests

GSG has received research grants and/or research support from the US National Institutes of Health, the University of Washington, the Bill & Melinda Gates Foundation, Gilead Sciences, Alere Technologies, Merck & Co., Janssen Pharmaceutica, Cerus Corporation, ViiV Healthcare, Bristol-Myers Squibb, Roche Molecular Systems, Abbott Molecular Diagnostics, and THERA Technologies/TaiMed Biologics, Inc, all outside of the submitted work. JAE reports grants to her institution from AstraZeneca, GlaxoSmithKline, Merck, and Pfizer and acts as a consultant for Abbvie, Ark Biopharma, AstraZeneca, GlaxoSmithKline, Meissa Vaccines, Merck, Moderna, Pfizer, and Sanofi Pasteur. MB has performed consulting for Allovir, Symbio, and Evrys Bio and has received research support from Merck. CML reports that her spouse is an employee of Bayer. HYC reports consulting for Ellume, Pfizer, and the Bill and Melinda Gates Foundation; has served on advisory boards for Vir, Merck and Abbvie; has conducted CME teaching with Medscape, Vindico, and Clinical Care Options; and has received research funding from Gates Ventures, and support and reagents from Ellume and Cepheid outside of the submitted work. All other authors have no competing interests to report.

## Supplements

### Supplementary Note S1

Nextstrain clades and Pango lineages are two of the most commonly used methods for classifying SARS-CoV-2. The Pango lineage system names lineages using a series of letters separated from a series of numbers by a period (for instance, A.1 and B.2.75). The system is hierarchical such that B.2.75 is a sublineage of the B.2 lineage. If a sublineage becomes common enough, it is assigned its own alphabetic name. For instance, B.1.1.529 is equivalent to BA. The Nextstrain clade system assigns names of two numbers corresponding to the year when the viral group was first observed and then a letter (for instance, 19A and 22C). In general, Nextstrain clades are usually larger (more inclusive) than Pango lineages, such that a clade will be composed of multiple lineages.

### Supplementary Note S2

For purposes of this study, we deemed an HCT participant to have been re-infected or superinfected with SARS-CoV-2 during the study period if they had at least two sequenced SARS-CoV-2 specimens that were determined to be of different clades and/or lineages. We did not observe any participants with 3 or more sequenced specimens each of different clades and/or lineages. It is possible that we excluded from the re-infected/superinfected participant group participants that experienced re-infection/superinfection with two closely related viruses (ie of the same clade and lineage). However, given the difficulty in distinguishing these situations from on-going shedding of the same virus, we chose to limit our list of re-infected/superinfected participants to those that had viruses of different clades and/or lineages.

### Supplementary Note S3

One of the 46 individuals identified as re-infected or superinfected had two sequenced specimens of two different clades/lineages collected only 6 days apart (first specimen is 22B/BA.5 and second is 21L/BA.2.3). The sequencing reads for both specimens were reviewed and there was no evidence in either specimen of mixed infection. These two specimens were the participant’s only SARS-CoV-2 positive specimens collected by HCT. Prior to testing positive the first time, the participant reported a known SARS-CoV-2 exposure but no recent travel. The participant denied having any history of previous SARS-CoV-2 infection prior to this first positive test. The participant reported experiencing symptoms for several days. The possibility of a specimen swap cannot be completely excluded, though there are numerous measures built into the study protocol to prevent such errors and a review of the documentation surrounding the collection of these specimens found no anomalies. Individuals testing using someone else’s information also cannot be excluded as a possibility, though is less likely in this case as both specimens were collected via staff observed in-person testing.

### Supplementary Figures

**Supplementary Figure S1:**
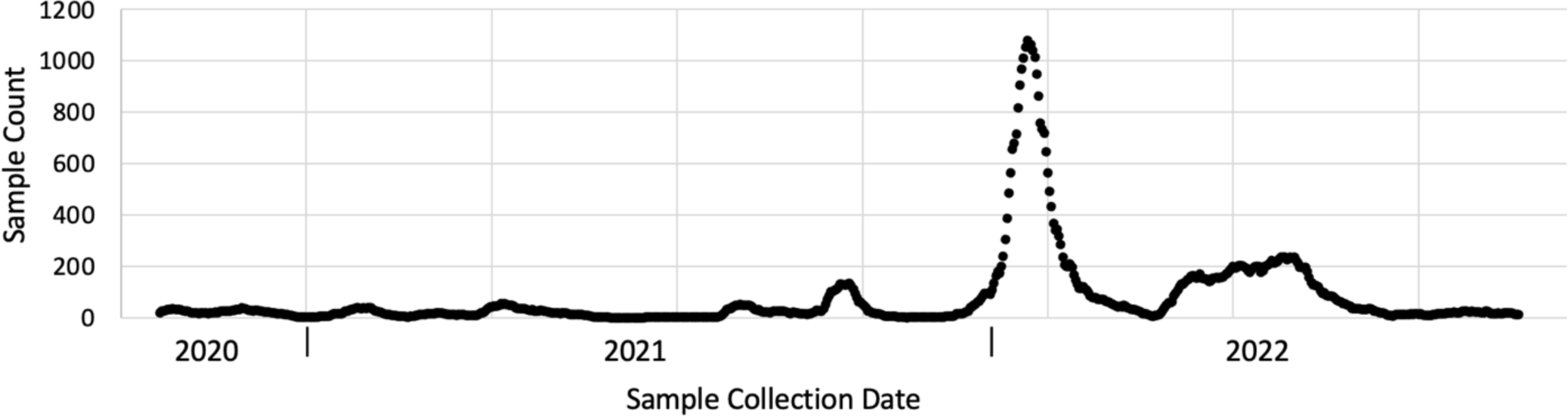
Total number of sequenced HCT specimens collected across the study period by two week sliding window.

**Supplementary Figure S2:**
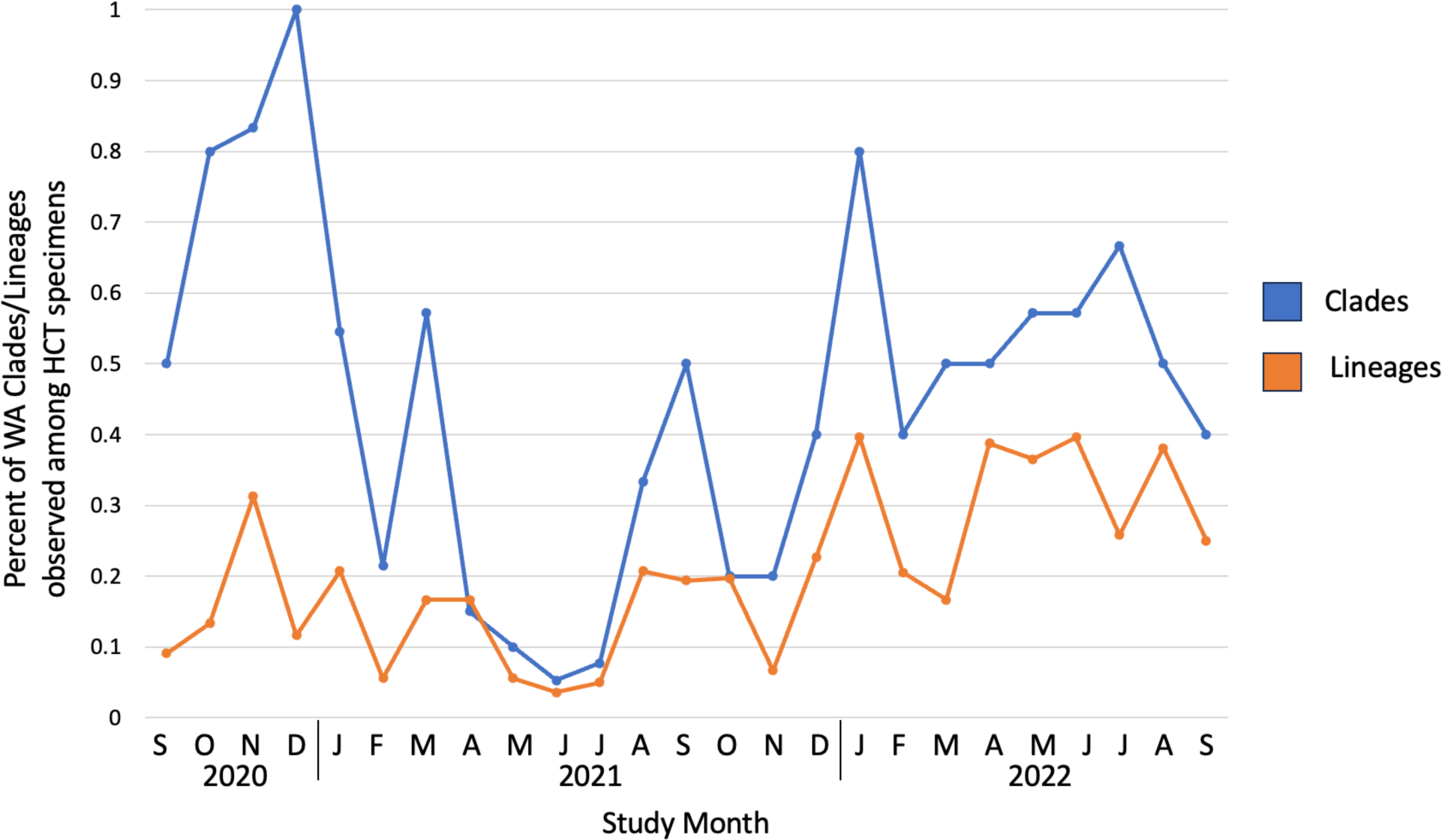
Percent of WA clades/lineages observed among sequenced HCT specimens by study month.

**Supplementary Figure S3:**
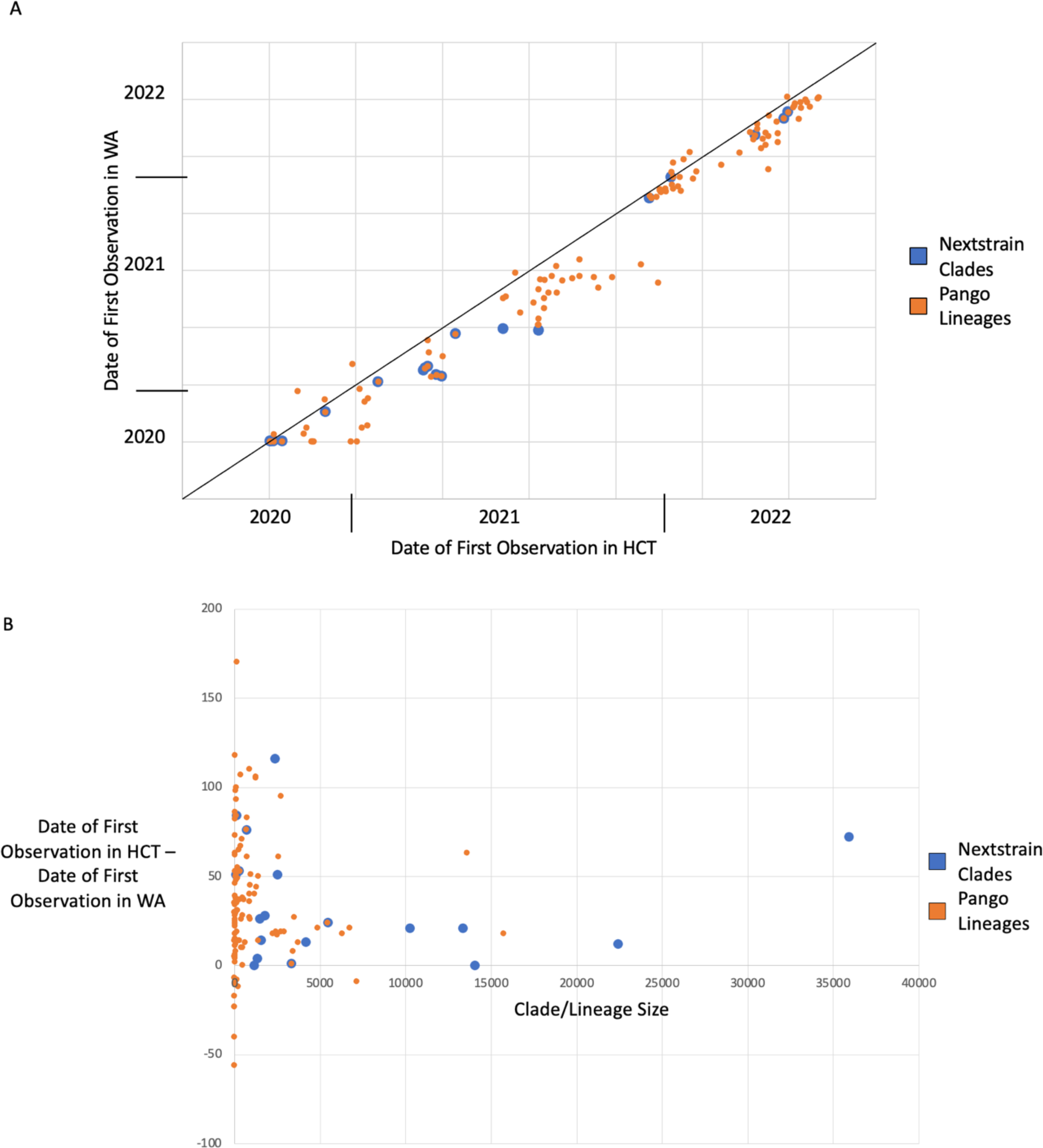
Date of first observation of clades and lineages in WA and in HCT. Blue dots represent Nextstrain clades. Orange dots represent Pango lineages. A) Chart with date of first observation in HCT on the x-axis and first observation in WA on the y-axis. B) Chart with size of clade/lineages on the x-axis and date of first observation in HCT minus first observation in WA (in days) on the y-axis.

**Supplementary Figure S4:**
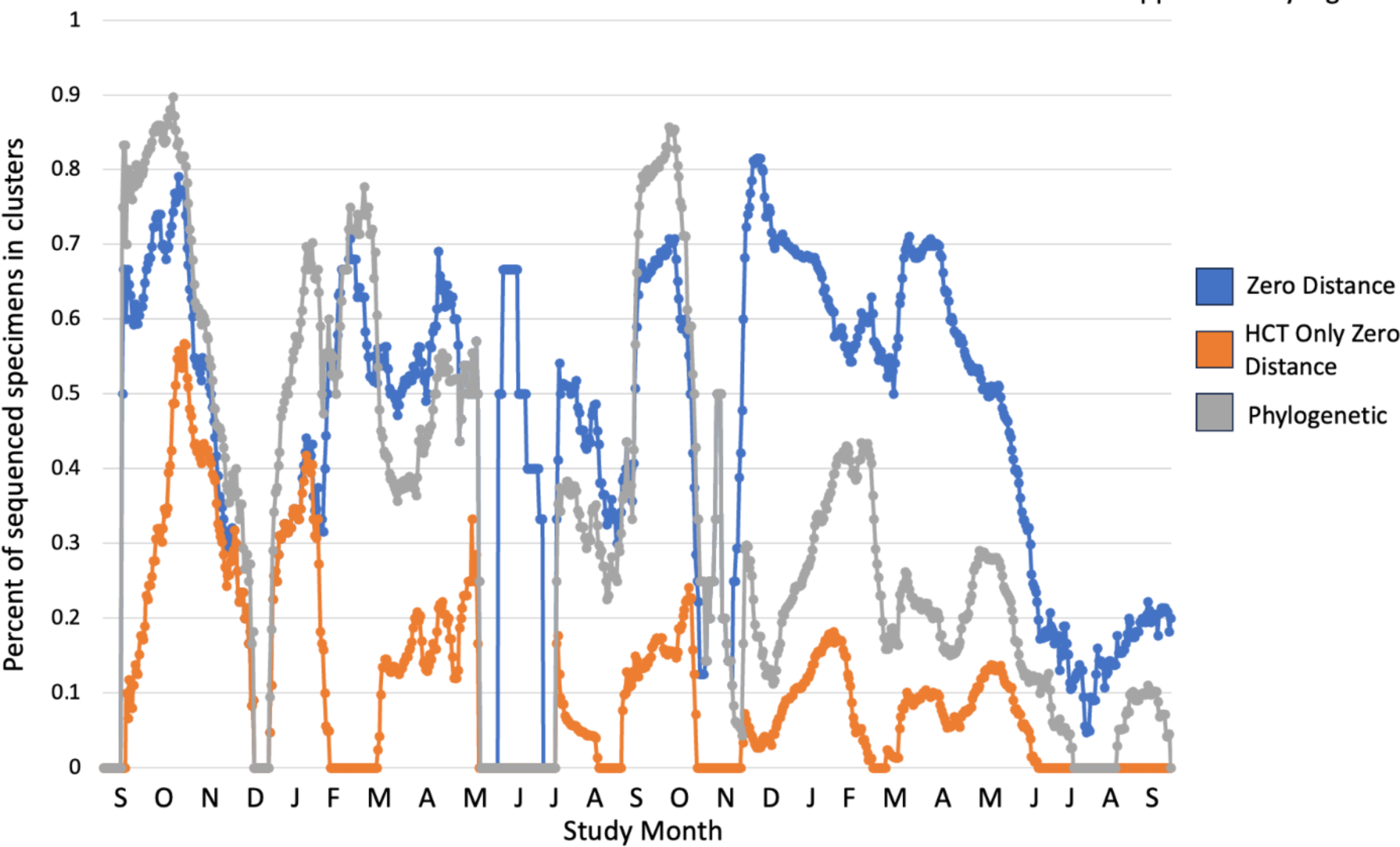
Percent of sequenced specimens in zero distance, HCT-only zero distance, and phylogenetic clusters calculated in one month sliding window periods.

**Supplementary Figure S5:**
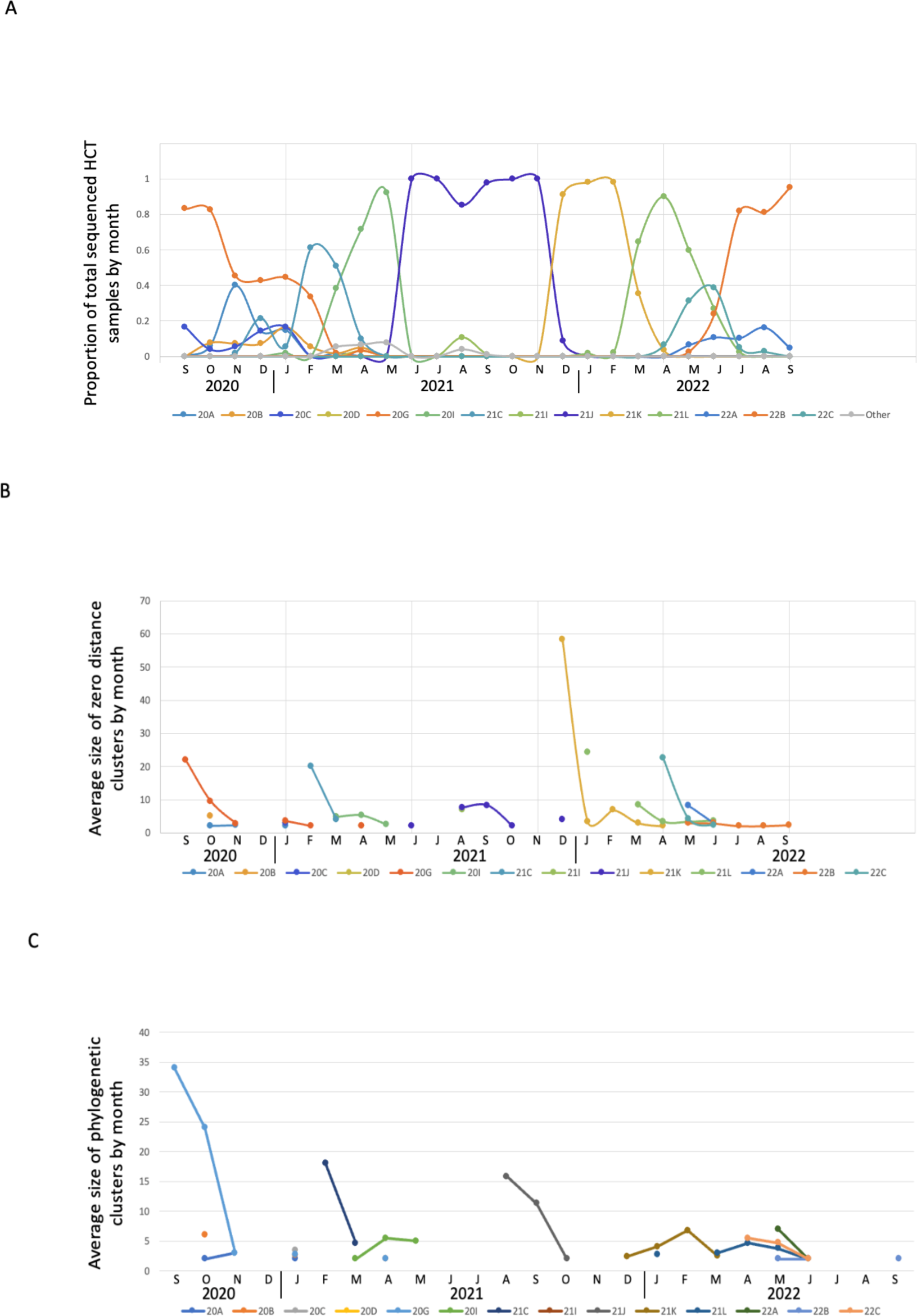
Average size of HCT sequence clusters by month. A) Proportion of total sequenced HCT specimens represented by each Nextstrain clade by month. B) Average size of zero distance clusters of each Nextstrain clade by month. C) Average size of phylogenetic clusters of each Nextstrain clade by month.

**Supplementary Figure S6:**
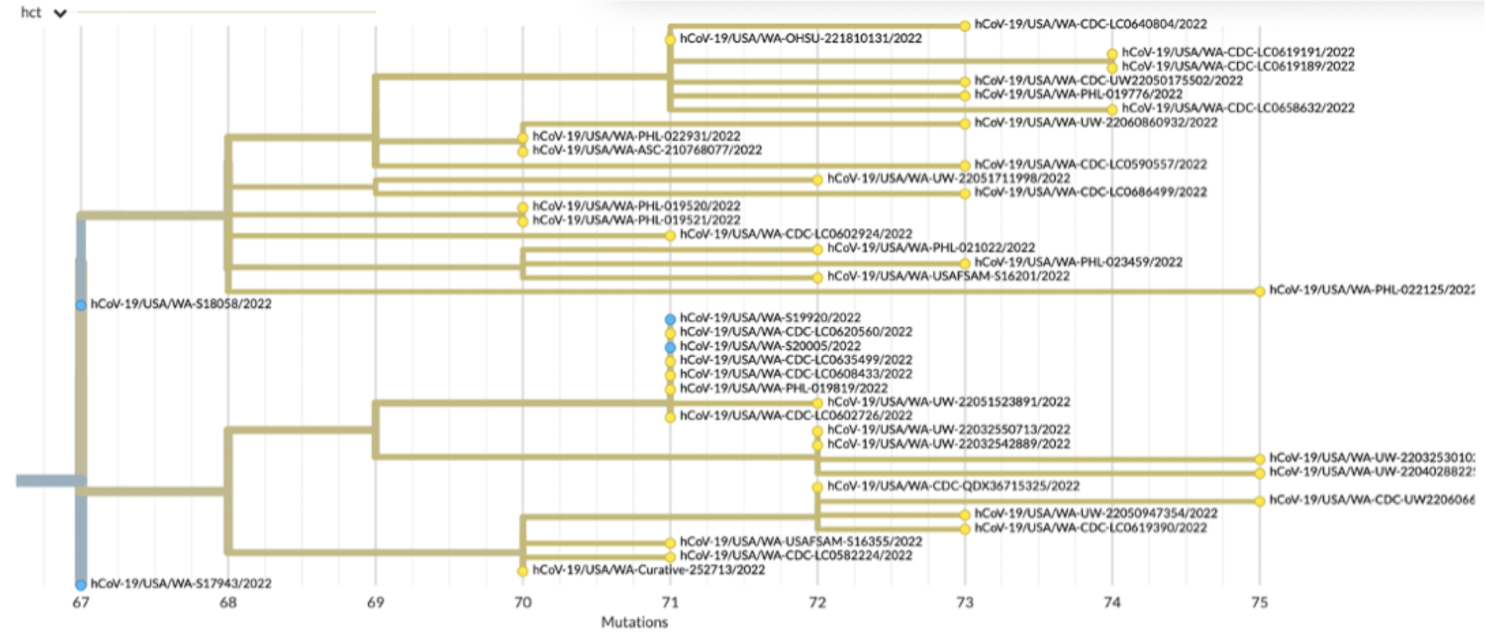
Phylogenetic tree showing HCT-only zero distance cluster with largest number of non-HCT descendants. HCT genomes are represented by blue nodes and non-HCT genomes by yellow nodes.

**Supplementary Figure S7:**
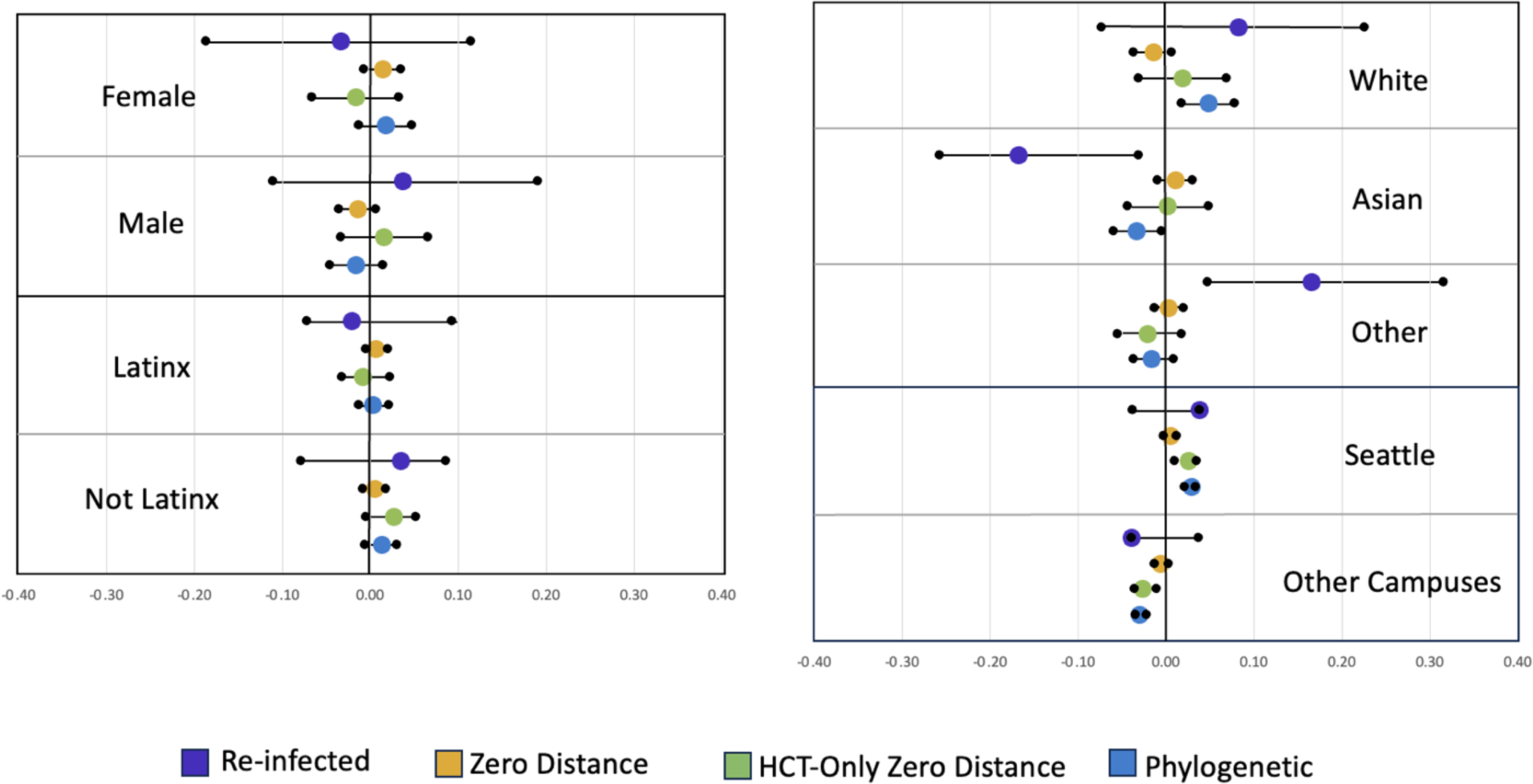
Demographic and epidemiologic characteristics of participants who experienced re-infection and of those with sequenced specimens in clusters. For each category (female, male, Latinx, Not Latinx, White, Asian, Other Race, Seattle campus, and other campuses), the mid-line (x = 0) represents the frequency of that category in the whole dataset. The purple, yellow, green, and blue dots give the percent difference between the frequency of that category among those who experienced re-infection and those with sequences in zero distance (groups of identical sequences), HCT-only zero distance (groups of identical sequences with haplotype unique to HCT), and phylogenetic clusters (groups of sequences that cluster phylogenetically), respectively, and the frequency of that category in the whole dataset. The intervals marked by black dots connected by a bar mark the 95% confidence interval of the values given by the purple, yellow, green, and blue dots.

## Notes

### Author Declarations

The IRB of the University of Washington gave ethical approval for this work.

